# Sporting tournaments and changed birth rates 9 months later: a systematic review

**DOI:** 10.1101/2022.12.12.22283373

**Authors:** Gwinyai Masukume, Victor Grech, Margaret Ryan

## Abstract

**Introduction:** Emerging evidence suggests that major sporting tournaments are associated with increased birth rates 9 months afterwards. This increase has been attributed to the celebratory atmosphere encouraging more conception through increased sexual activity among the populace.

**Methods:** Studies that examined the relationship between a major sporting tournament and altered birth metrics (number of births and/or birth sex ratio, defined as male divided by total live births) 9(±1) months later were sought out systematically through searches of PubMed and Scopus. Preferred Reporting Items for Systematic Reviews and Meta-Analyses guidelines were followed in conducting this study.

**Results:** Five major sporting tournaments were linked to noticeably increased birth metrics 9(±1) months later. The Super Bowl (increased United States birth sex ratio in multiple years, however, from 2004 to 2013 there were no observable birth increases in winning counties and losing was not associated with a changed number of births), the 2009 UEFA Champions League (16% increase in Solsonès and Bages births in Spain), the 2010 FIFA World Cup (increased birth sex ratio and over 1000 extra births in South Africa), the 2016 UEFA Euros (2% increase in Northern Ireland births) and the 2019 Rugby World Cup (increased birth sex ratio in some Japanese prefectures). Nine months after the most popular provincial *la Liga* soccer teams unexpectedly lost matches, there were 0.8% fewer births in those provinces from 2001 to 2015; the number of births were unaffected by unexpected wins. After the 1998 FIFA World Cup a changed sex ratio at birth was not witnessed 9 months on. Nine months following a performance improvement of one standard deviation by a European national soccer team at the FIFA World Cup or UEFA Euro Championships, from 1960 to 2016, there was a 0.3% decline in births 9 months on.

**Conclusions:** With a few exceptions, major American football, Association football (soccer) and Rugby apex tournaments in Africa, North America, Asia and Europe were associated with increases in the number of babies born and/or in the birth sex ratio 9(±1) months following notable team wins and/or hosting the tournament. Related to this, unexpected losses by teams from a premier soccer league were associated with a decline in births 9 months on. In conclusion, Nelson Mandela was correct when he averred, “Sport has the power to change the world. It has the power to inspire, it has the power to unite people in a way that little else does.”

**PROSPERO registration:** CRD42022382971

## Introduction

A rise in the number of births 9 months later has been linked, in some settings, to a variety of events, including carnivals, religious and secular holidays such as Christmas and New Year’s Day (January 1).^1-3^ This increase has been attributed to the holiday celebrations encouraging more conception, 9 months earlier,^4^ through increased sexual activity among the populace.

Sports are a dominant part of culture around the world, with major sporting tournaments attracting millions of attendees and even billions of viewers.^5^ It is important to consider how the outcomes of these tournaments affect population-level demographics because fans often have a deep investment in the triumph of their beloved teams which may influence fertility.^6^ Emerging evidence suggests that major sporting tournaments are also associated with increased birth rates 9 months afterwards. For instance, in Northern Ireland (NI), 9 months after the 2016 Union of European Football Associations (UEFA) Euros Championship, in which NI was competing for the first time, a 2% rise in births was observed in March 2017.^7^ Nine months following Futbol Club Barcelona’s victory in the 2009 UEFA Champion’s league, there was a considerable 16% increase in births in Catalonia.^8^ On the other hand, it has been reported that 9 months following the United States (US) National Football’s Super Bowl, the number of births in winning cities did not rise.^9^ However, the US sex ratio at birth (SRB), defined as male divided by total live births, increased 9 months after several Super Bowls.^10^ Nine months after the 2010 *Fédération Internationale de Football Association* (FIFA) World Cup, there were over 1000 more live births, and the SRB was higher.^11, 12^ In addition, there have been several anecdotal media stories about baby booms 9 months after sporting tournaments, for instance a boom in Iceland after the 2016 UEFA Euros Championship, but subsequent statistical analyses of birth data has not supported these claims.^13^ In contrast to the aforementioned, but consistent with the paradigm, unexpected losses of Spanish local soccer teams have been reported to lead to a 0.8% decrease in the local number of births 9 months later.^14^

Given the sometimes conflicting and inconsistent reports, our objective was to conduct a systematic review of the literature evaluating the relationship between sporting events and live birth rate/proportion changes 9(±1) months later. If sporting events alter birth patterns significantly, this could have an impact on the need for midwifery, medical and other healthcare personnel as well as resources.

## Methods

The protocol for this systematic review adheres to the Preferred Reporting Items for Systematic Reviews and Meta-Analyses (PRISMA) standards.^15^ This review was registered on the International prospective register of systematic reviews site PROSPERO, CRD42022382971.

### Search strategy

A search strategy and search terms were developed for this study. Without regard to language, a systematic literature search was conducted. Using the search strategy (Table 1), potentially relevant studies were looked up from database inception through to 7 November 2022 in the electronic databases PubMed and Scopus. The first author carried out all searches. The following search term syntax was used:

**Table 1.** Search terms used to identify relevant studies.

| Exposure | Outcome |
| --- | --- |
| PubMed |  |
| MeSH term “Sports” | MeSH term “Birth rate” |
| Athletic Performance | Birth Rates |
| Baseball | Rate, Birth |
| Basketball | Natality |
| Bicycling | Natalities |
| Boxing | Fertility Rate |
| Cricket Sport | Fertility Rates |
| Football | Rate, Fertility |
| Golf | Age-Specific Birth Rate |
| Gymnastics | Age Specific Birth Rate |
| Hockey | Age-Specific Birth Rates |
| Martial Arts | Birth Rate, Age-Specific |
| Mountaineering | Age-Specific Fertility Rate |
| Racquet Sports | Age Specific Fertility Rate |
| Return to Sport | Age-Specific Fertility Rates |
| Rugby | Fertility Rate, Age-Specific |
| Running | Rate, Age-Specific Fertility |
| Skating | MeSH term “Sex Ratio” |
| Snow Sports | Ratio, Sex |
| Soccer | Ratios, Sex |
| Sports for Persons with Disabilities | Sex Ratios |
| Team Sports | MeSH term “Fertility” |
| Track and Field | Fecundability |
| Volleyball | Fecundity |
| Walking | Differential Fertility |
| Water Sports | Fertility, Differential |
| Weight Lifting | Fertility Determinants |
| Wrestling | Determinant, Fertility |
| Youth Sports | Determinants, Fertility |
|  | Fertility Determinant |
|  | Subfecundity |
|  | Fertility Preferences |
|  | Fertility Preference |
|  | Preference, Fertility |
|  | Preferences, Fertility |
|  | Fertility, Below Replacement |
|  | Below Replacement Fertility |
|  | Marital Fertility |
|  | Fertility, Marital |
|  | Natural Fertility |
|  | Fertility, Natural |
|  | World Fertility Survey |
|  | Fertility Survey, World |
|  | Fertility Surveys, World |
|  | Survey, World Fertility |
|  | Surveys, World Fertility |
|  | World Fertility Surveys<br>Fertility Incentives<br>Fertility Incentive |
| Scopus |  |
| KEY field “Sports” | KEY field “births”<br>KEY field “birth rate”<br>KEY field “fertility” |

MeSH – Medical Subject Heading. The Scopus Field code KEY was used in an Advanced search. The KEY field code is a combined code which searches AUTHKEY, INDEXTERM, TRADENAME and CHEMNAME fields. AUTHKEY (Key words assigned to the document by the author), INDEXTERM (controlled vocabulary terms assigned to the document), TRADENAME (a name used to identify a commercial product or service) and CHEMNAME (Chemical Name).

PubMed: *“Sports”[Mesh] AND (“Birth Rate”[Mesh] OR “Sex ratio”[MESH] OR “Fertility”[MESH])*

Scopus: *KEY (“sports”) AND KEY (“birth”) OR KEY (“birth rate”) OR KEY (“fertility”)*

Using Covidence systematic review software (www.covidence.org), we completed the title/abstract and full-text screening for this study. This was performed independently by two authors (GM and MR). The reference lists of the studies that were included in the search were manually searched as an add-on. The ‘cited by’ function of Google Scholar was also used to find articles citing the included studies.

### Eligibility

Table 2 lists the inclusion and exclusion criteria that were employed in this study.

**Table 2.**
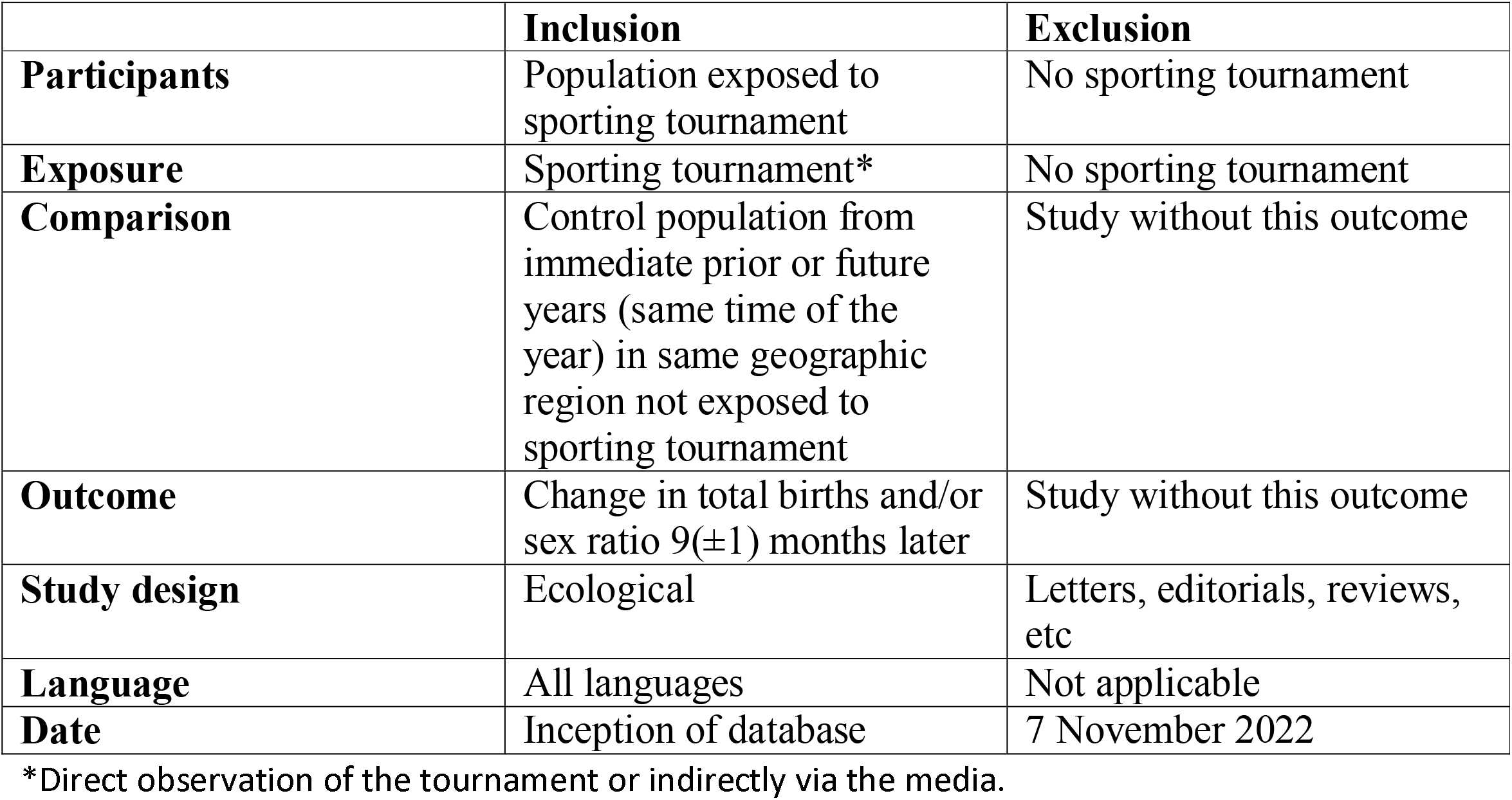
Inclusion and exclusion criteria.

|  | **Inclusion** | **Exclusion** |
| --- | --- | --- |
| **Participants** | Population exposed to sporting tournament | No sporting tournament |
| **Exposure** | Sporting tournament* | No sporting tournament |
| **Comparison** | Control population from immediate prior or future years (same time of the year) in same geographic region not exposed to sporting tournament | Study without this outcome |
| **Outcome** | Change in total births and/or sex ratio 9(±1) months later | Study without this outcome |
| **Study design** | Ecological | Letters, editorials, reviews, etc |
| **Language** | All languages | Not applicable |
| **Date** | Inception of database | 7 November 2022 |

\*Direct observation of the tournament or indirectly via the media.

### Data extraction

Data were extracted from the included studies by two reviewers (GM and MR). Author name and year, country or region of the tournament, sport and change in birth pattern (total births/SRB) were extracted. Extraction differences were discussed by these two authors and were resolved.

### Quality assessment

As far as we are aware, there is no instrument that has been validated for use in evaluating the quality of ecological studies.^16^ However, the group exposure (sporting tournament) was assessed according to how the population that experienced the tournament defined it; in other words, there was common understanding that the sporting tournament was significant, and the studies characterized it as such. There was minimal concern about outcome bias because the human live birth statistics of the entire population (and not just a sample) was available and the danger of misclassifying sex at birth is low.^17^

## Results

The search strategy was used to find 111 and 332 articles from PubMed and Scopus respectively. After uploading the search results into Covidence and removing 12 duplicates the authors (GM and MR) evaluated each of the remaining 431 articles’ titles and abstracts individually (Figure 1). After examining all titles and abstracts independently, these authors convened to discuss and resolve any differences. All differences were resolved. Six articles remained (Figure 1). An additional 4 articles were found by citation searching. The final data extraction included all 10 of the articles that made it to the full-text screening stage. The characteristics of sporting events with altered birth rates or SRB 9(±1) months later are listed in Table 3. Major American football, Association football (soccer) and rugby championships were linked to increases in the number of births and/or birth sex ratio 9(±1) months after notable team victories and/or the tournament’s hosting. Related to this, a drop in birth rates 9 months later was linked to a team from a top soccer league suffering unexpected losses (Table 3).

**Table 3.**
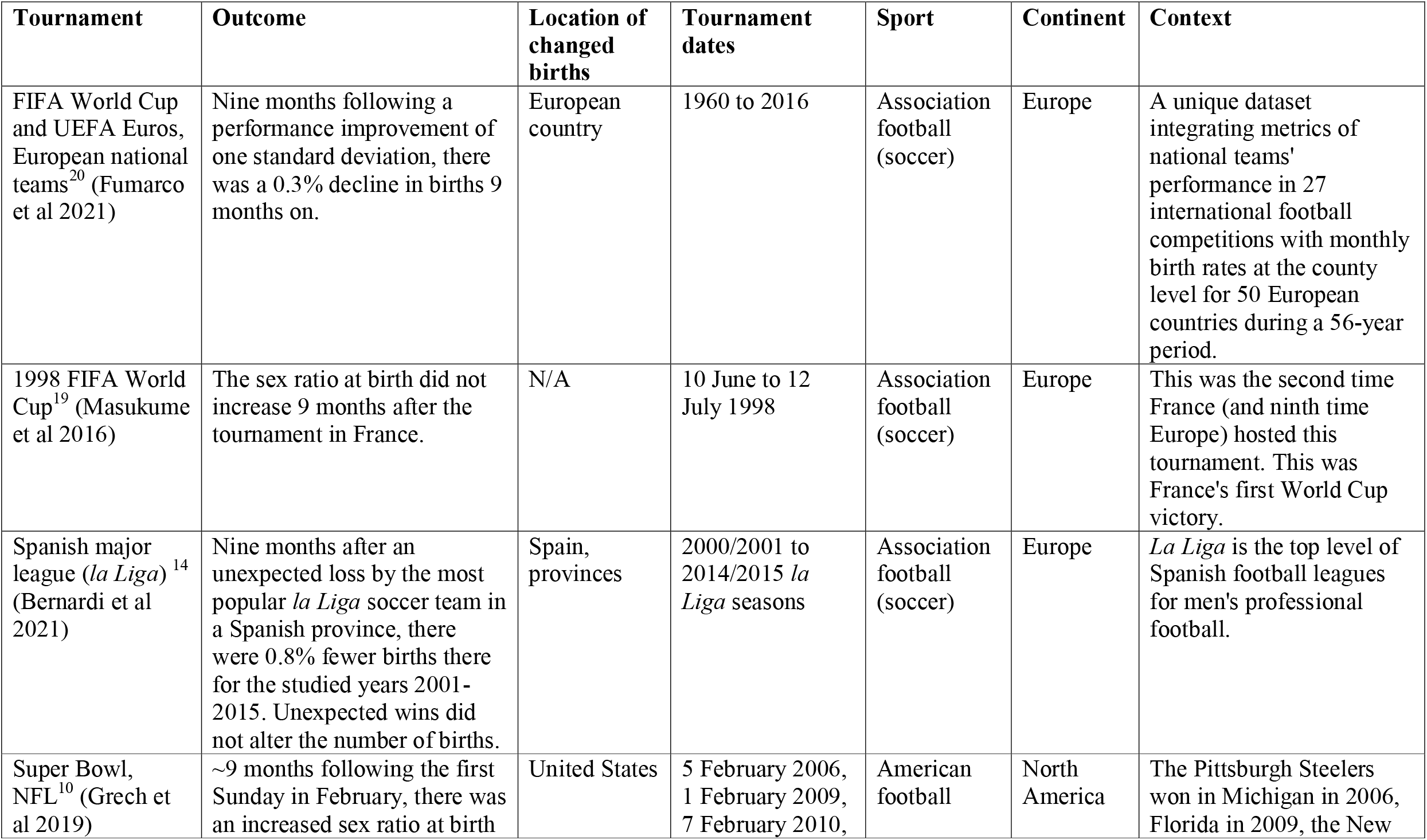
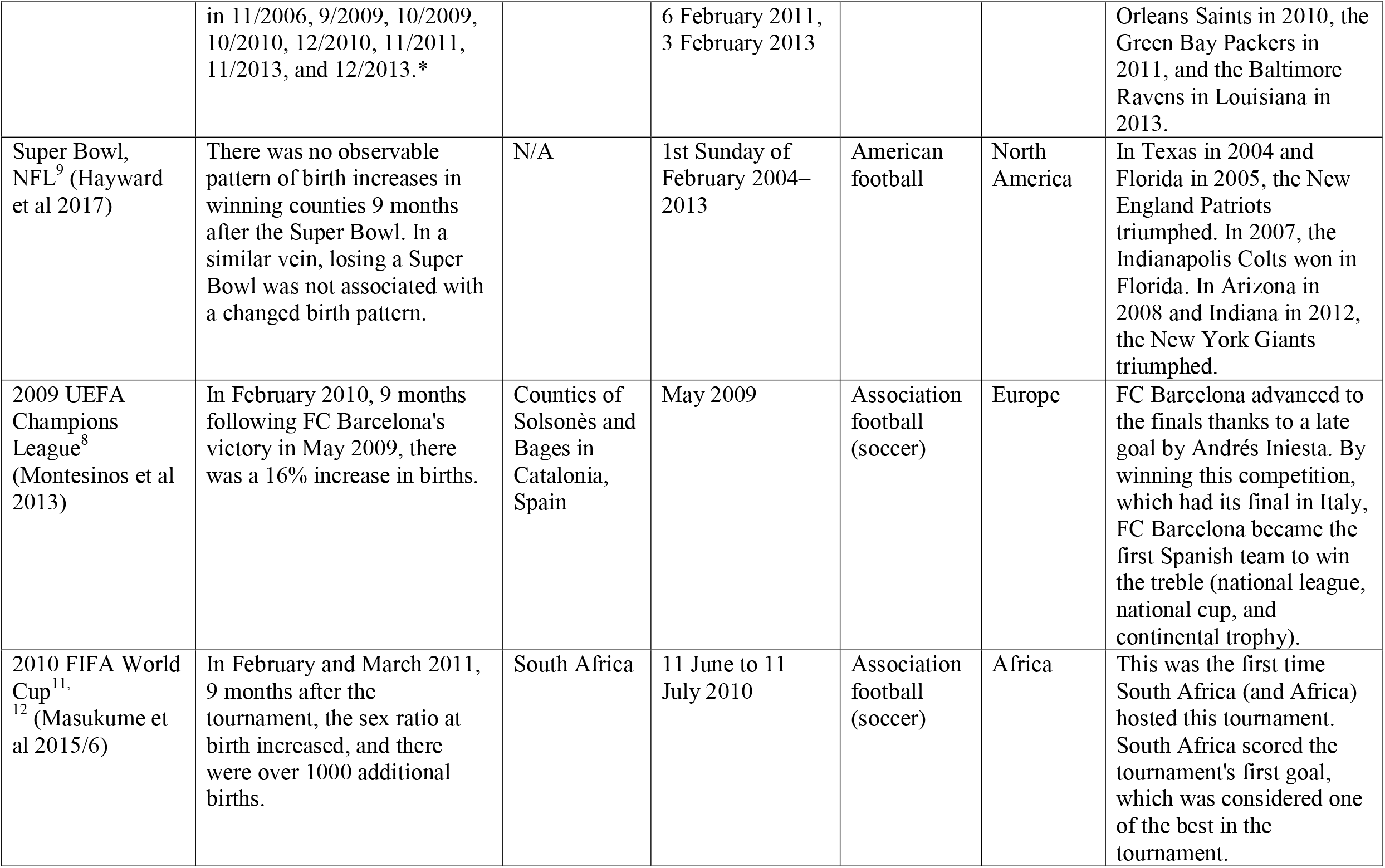
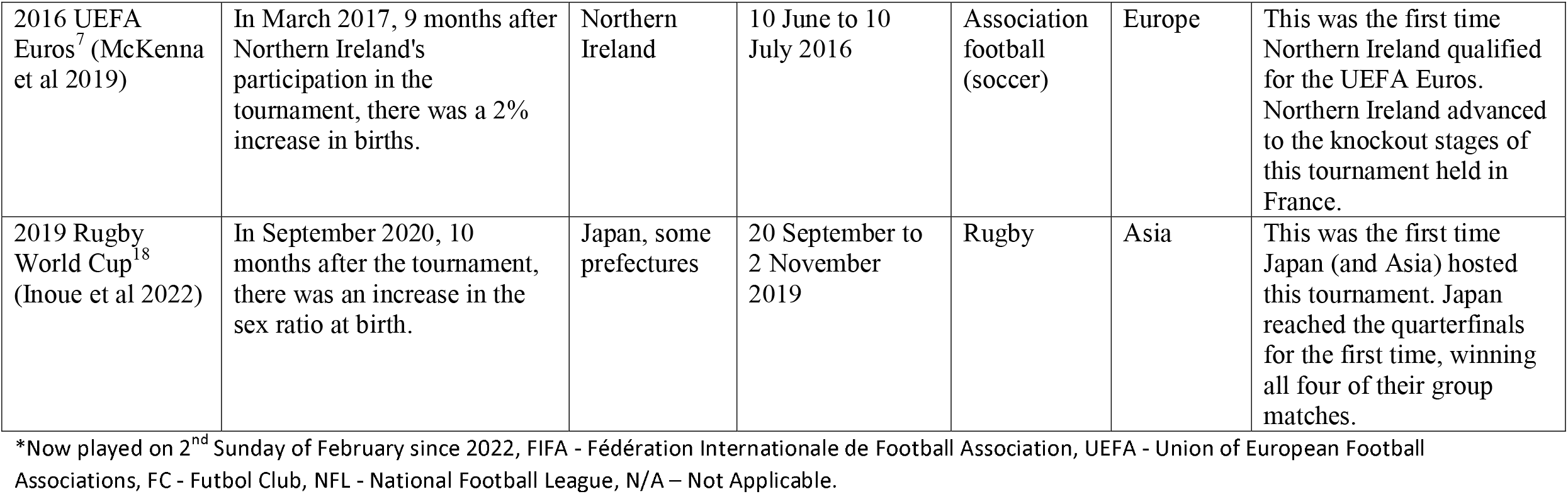
Characteristics of included sporting tournaments.

| **Tournament** | **Outcome** | **Location of changed births** | **Tournament dates** | **Sport** | **Continent** | **Context** |
| --- | --- | --- | --- | --- | --- | --- |
| FIFA World Cup and UEFA Euros, European national teams<sup>20</sup> (Fumarco et al 2021) | Nine months following a performance improvement of one standard deviation, there was a 0.3% decline in births 9 months on. | European country | 1960 to 2016 | Association football (soccer) | Europe | A unique dataset integrating metrics of national teams' performance in 27 international football competitions with monthly birth rates at the county level for 50 European countries during a 56-year period. |
| 1998 FIFA World Cup<sup>19</sup> (Masukume et al 2016) | The sex ratio at birth did not increase 9 months after the tournament in France. | N/A | 10 June to 12 July 1998 | Association football (soccer) | Europe | This was the second time France (and ninth time Europe) hosted this tournament. This was France's first World Cup victory. |
| Spanish major league (*la Liga*)<sup>14</sup> (Bernardi et al 2021) | Nine months after an unexpected loss by the most popular *la Liga* soccer team in a Spanish province, there were 0.8% fewer births there for the studied years 2001-2015. Unexpected wins did not alter the number of births. | Spain, provinces | 2000/2001 to 2014/2015 *la Liga* seasons | Association football (soccer) | Europe | *La Liga* is the top level of Spanish football leagues for men's professional football. |
| Super Bowl, NFL<sup>10</sup> (Grech et al 2019) | ~9 months following the first Sunday in February, there was an increased sex ratio at birth | United States | 5 February 2006, 1 February 2009, 7 February 2010, | American football | North America | The Pittsburgh Steelers won in Michigan in 2006, Florida in 2009, the New |
|  | in 11/2006, 9/2009, 10/2009, 10/2010, 12/2010, 11/2011, 11/2013, and 12/2013.* |  | 6 February 2011, 3 February 2013 |  |  | Orleans Saints in 2010, the Green Bay Packers in 2011, and the Baltimore Ravens in Louisiana in 2013. |
| Super Bowl, NFL<sup>9</sup> (Hayward et al 2017) | There was no observable pattern of birth increases in winning counties 9 months after the Super Bowl. In a similar vein, losing a Super Bowl was not associated with a changed birth pattern. | N/A | 1st Sunday of February 2004–2013 | American football | North America | In Texas in 2004 and Florida in 2005, the New England Patriots triumphed. In 2007, the Indianapolis Colts won in Florida. In Arizona in 2008 and Indiana in 2012, the New York Giants triumphed. |
| 2009 UEFA Champions League<sup>8</sup> (Montesinos et al 2013) | In February 2010, 9 months following FC Barcelona's victory in May 2009, there was a 16% increase in births. | Counties of Solsonès and Bages in Catalonia, Spain | May 2009 | Association football (soccer) | Europe | FC Barcelona advanced to the finals thanks to a late goal by Andrés Iniesta. By winning this competition, which had its final in Italy, FC Barcelona became the first Spanish team to win the treble (national league, national cup, and continental trophy). |
| 2010 FIFA World Cup<sup>11, 12</sup> (Masukume et al 2015/6) | In February and March 2011, 9 months after the tournament, the sex ratio at birth increased, and there were over 1000 additional births. | South Africa | 11 June to 11 July 2010 | Association football (soccer) | Africa | This was the first time South Africa (and Africa) hosted this tournament. South Africa scored the tournament's first goal, which was considered one of the best in the tournament. |
| 2016 UEFA Euros<sup>7</sup> (McKenna et al 2019) | In March 2017, 9 months after Northern Ireland's participation in the tournament, there was a 2% increase in births. | Northern Ireland | 10 June to 10 July 2016 | Association football (soccer) | Europe | This was the first time Northern Ireland qualified for the UEFA Euros. Northern Ireland advanced to the knockout stages of this tournament held in France. |
| 2019 Rugby World Cup<sup>18</sup> (Inoue et al 2022) | In September 2020, 10 months after the tournament, there was an increase in the sex ratio at birth. | Japan, some prefectures | 20 September to 2 November 2019 | Rugby | Asia | This was the first time Japan (and Asia) hosted this tournament. Japan reached the quarterfinals for the first time, winning all four of their group matches. |

\*Now played on 2<sup>nd</sup> Sunday of February since 2022, FIFA - Fédération Internationale de Football Association, UEFA - Union of European Football Associations, FC - Futbol Club, NFL - National Football League, N/A – Not Applicable.

**Figure 1.**
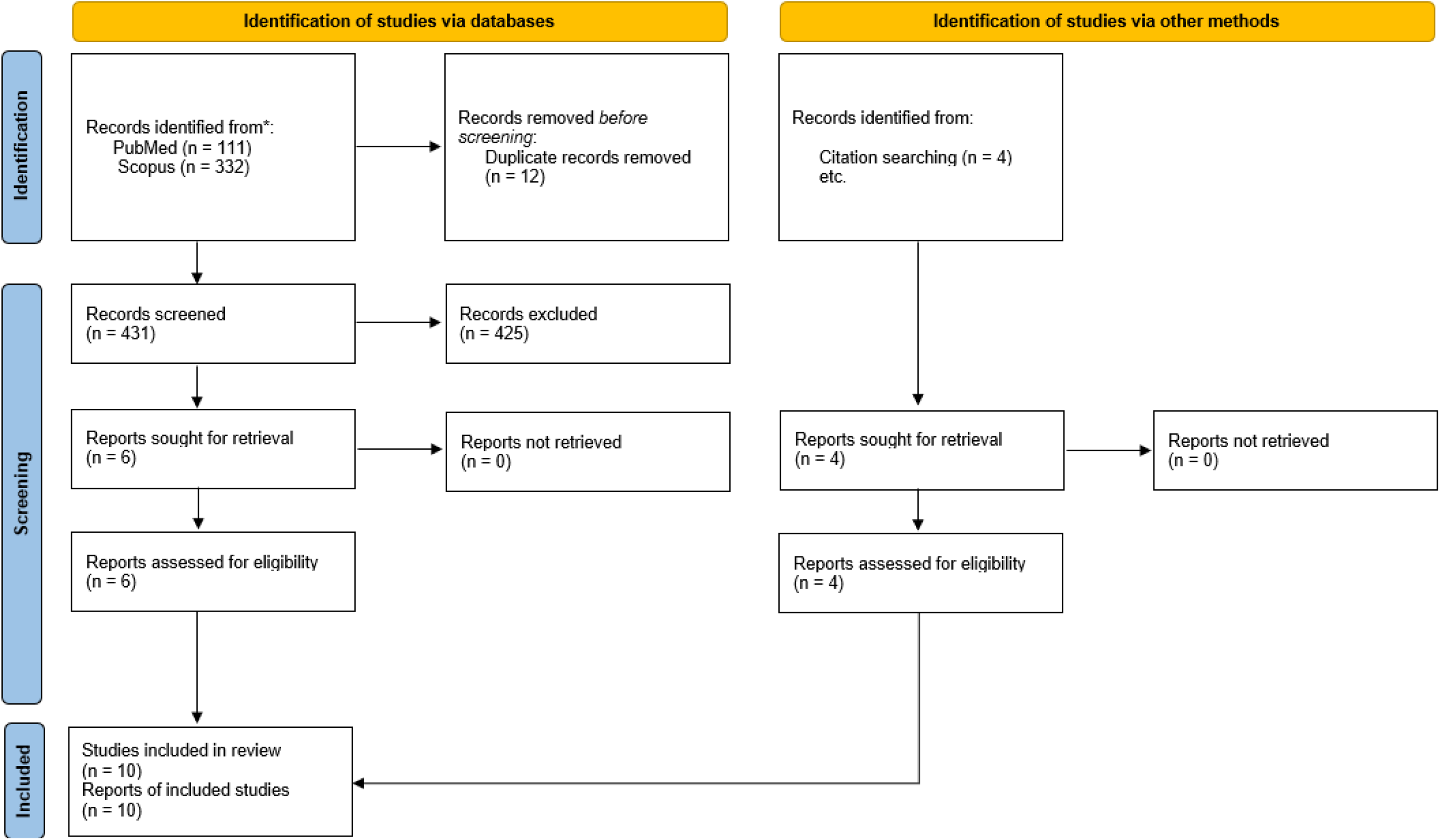
Flow chart of studies identified for inclusion.

## Discussion

### Principle findings

In this study, we found that five major sporting tournaments were linked to noticeably increased birth metrics 9(±1) months later. The Super Bowl (increased US birth sex ratio in multiple years,^10^ however, from 2004 to 2013 there were no observable birth increases in winning counties and losing was not associated with a changed number of births^9^), the 2009 UEFA Champions League (16% increase in Solsonès and Bages births in Spain^8^), the 2010 FIFA World Cup (increased birth sex ratio^11^ and over 1000 extra births in South Africa^12^), the 2016 UEFA Euros (2% increase in Northern Ireland births^7^) and the 2019 Rugby World Cup (increased birth sex ratio in some Japanese prefectures^18^). Nine months after the most popular provincial *la Liga* soccer teams unexpectedly lost matches, there were 0.8% fewer births in those provinces from 2001 to 2015; the number of births were unaffected by unexpected wins.^14^ After the 1998 FIFA World Cup a changed sex ratio at birth was not witnessed 9 months on.^19^ Nine months following a performance improvement of one standard deviation by a European national soccer team at the FIFA World Cup or UEFA Euro Championships, from 1960 to 2016, there was a 0.3% decline in births 9 months on.^20^

### Comparison with other studies

Some studies have linked a surge in births 9 months later to several occasions, such as carnivals, religious events and secular holidays like Christmas and New Year’s Day (January 1). This increase has been attributed to the holiday celebrations, which promoted greater conception through increased sexual activity among the public.^1-4^ Although exceptions exist, the principle finding of this systematic review, of baby booms 9(±1) months after winning or hosting major sports tournaments, across different sports and continents, is consistent with the mechanism of celebratory sexual intercourse.^9^ After a team wins, it has been hypothesized that a person’s happiness coincides with a hormonal shift that elevates their desire for sexual activity.^21^ Large-scale human emotions can therefore have a significant impact on population demographic changes.

Despite the fact that there was no general increase in live births after the Super Bowl,^9^ the ratio of male to female live births increased nine months later.^10^ While both the number of births and the male to female live birth ratio increased after the 2010 FIFA World Cup. This indicates that it is useful to consider both the overall number of live births and the births’ male to female ratio (SRB).

Although England, Northern Ireland, Scotland and Wales are separate countries with their own national soccer teams, the United Kingdom was treated as a single entity in one investigation of this phenomenon.^20^ According to this investigation, there was a 0.3% drop in births 9 months after a national team performance increase of one standard deviation. The 2016 UEFA tournament in which Northern Ireland participated, had a 2% increase in NI births 9 months later, which appears to be in conflict with this investigation.^7^ The findings of this investigation^20^ should thus be handled cautiously given how the United Kingdom was treated as a single nation.

The context in which baby booms take place appears to be important, such as when a nation hosts big international sporting events for the first time (e.g. South Africa 2010 FIFA World Cup,^11, 12^ Japan 2019 Rugby World Cup^18^), or when it first qualifies for one and does relatively well (Northern Ireland 2016 UEFA Euros^7^). It is conceivable that during these inaugural tournaments, public excitement is at its peak, increasing the frequency of celebratory sexual intercourse. This may help to explain why France, despite winning the tournament, did not experience a baby boom 9 months after the 1998 FIFA World Cup as it was the country’s second time hosting the tournament and it had previously qualified.^19^ Unexpected defeats in the Spanish top division *la Liga*, when fewer births were recorded 9 months later, are another contextual element.^14^ An unexpected defeat is more likely to have a negative impact on mood than an expected defeat, with probable aftereffects including a reduction in the number of sexual encounters and subsequent births 9 months later.

That a change in the number of babies born or in the sex ratio at birth occurred circa 9 months after major sporting tournaments across different sports and continents suggests a true effect and a common underlying mechanism. This mechanism suggests that euphoria or sadness experienced when a sports team wins or loses coincides with a hormonal change that increases or decreases a person’s desire for sexual activity, which then affects birth rates nine months later.

### Strengths and limitations

A noteworthy strength is the methodology, which is transparent and replicable, and does not impose any language restrictions on the search. Our investigation was constrained because we could have overlooked important papers by not searching gray literature.

This study has the same drawbacks as ecological studies, such as its restricted capacity to draw conclusions at the individual level (the ecological fallacy).^22^ However, it is probable that ecological research is one of the best ways to assess how a population event affects a population outcome.^23^

## Conclusions

With a few exceptions, major American football, Association football (soccer) and Rugby apex tournaments in Africa, North America, Asia and Europe were associated with increases in the number of babies born and/or in the birth sex ratio 9(±1) months following notable team wins and/or hosting the tournament. Related to this, unexpected losses by teams from a premier soccer league were associated with a decline in births 9 months on. The results of this systematic review suggest that sporting events considerably modify birth patterns, which may affect the demand for midwives, doctors, and other healthcare staff as well as resources.

In conclusion, Nelson Mandela was correct when he averred, “Sport has the power to change the world. It has the power to inspire, it has the power to unite people in a way that little else does.”^24^

## Data Availability

All data produced in the present work are contained in the manuscript.

## Ethics

Not required as a systematic review of published data.

## References

1. Kadhel P, Costet N, Toto T, Janky E, Multigner L. The annual carnival in Guadeloupe (French West Indies) is associated with an increase in the number of conceptions and subsequent births nine months later: 2000 – 2011. PLOS ONE. 2017;12(3):e0173102.

2. Macfarlane A, Dattani N, Gibson R, Harper G, Martin P, Scanlon M, et al. Births and their outcomes by time, day and year: a retrospective birth cohort data linkage study. Health Services and Delivery Research. 2019;7(18).

3. Régnier-Loilier A, Rohrbasser J-M. Y a-t-il une saison pour faire des enfants? Population Societes. 2011(1):1–4.

4. Jukic AM, Baird DD, Weinberg CR, McConnaughey DR, Wilcox AJ. Length of human pregnancy and contributors to its natural variation. Hum Reprod. 2013;28(10):2848–55.

5. Maennig W, Zimbalist A. Introduction: The economics of mega sporting events. International handbook on the economics of mega sporting events: Edward Elgar Publishing; 2012.

6. Gorelik G, Bjorklund DF. The Effect of Competition on Men’s Self-Reported Sexual Interest. Evolutionary Psychological Science. 2015;1(3):141–9.

7. McKenna CS, Znaczko A, Morrison PJ. BIRTH RATE MAY INCREASE NINE MONTHS AFTER NATIONAL FOOTBALL SUCCESS. Ulster Med J. 2019;88(1):56–8.

8. Montesinos J, Cortes J, Arnau A, Sanchez JA, Elmore M, Macia N, et al. Barcelona baby boom: does sporting success affect birth rate? BMJ : British Medical Journal. 2013;347:f7387.

9. Hayward GM, Rybińska A. “Super Bowl Babies”: Do Counties with Super Bowl Winning Teams Experience Increases in Births Nine Months Later? Socius. 2017;3.

10. Grech V, Zammit D. Influence of the Super Bowl on the United States birth sex ratio. Early Hum Dev. 2019;128:86–92.

11. Masukume G, Grech V. The sex ratio at birth in South Africa increased 9months after the 2010 FIFA World Cup. Early Hum Dev. 2015;91(12):807–9.

12. Masukume G, Grech V, Scherb H. Reply: Analysis of the sex ratio at birth in South Africa increased 9months after the 2010 FIFA World Cup. Early Hum Dev. 2016;97:11–3.

13. Grech V, Masukume G. Fake news of baby booms 9months after major sporting events distorts the public’s understanding of early human development science. Early Hum Dev. 2017;115:16–7.

14. Bernardi F, Cozzani M. Soccer Scores, Short-Term Mood and Fertility. Eur J Popul. 2021;37(3):625–41.

15. Page MJ, Moher D, Bossuyt PM, Boutron I, Hoffmann TC, Mulrow CD, et al. PRISMA 2020 explanation and elaboration: updated guidance and exemplars for reporting systematic reviews. BMJ. 2021;372:n160.

16. Betran AP, Torloni MR, Zhang J, Ye J, Mikolajczyk R, Deneux-Tharaux C, et al. What is the optimal rate of caesarean section at population level? A systematic review of ecologic studies. Reprod Health. 2015;12:57.

17. Davis DL, Gottlieb MB, Stampnitzky JR. Reduced ratio of male to female births in several industrial countries: a sentinel health indicator? Jama. 1998;279(13):1018–23.

18. Inoue Y, Mizoue T. A preliminary analysis of the secondary sex ratio decline after the COVID-19 pandemic in Japan. Am J Hum Biol. 2022:eajhb.

19. Masukume G, Grech V. The sex ratio at birth in France was unchanged 9months after the 1998 FIFA World Cup. Early Hum Dev. 2016;99:13–5.

20. Fumarco L, Principe F. More goals, fewer babies? On national team performance and birth rates. Economics Letters. 2021;208:110086.

21. Casto KV, Edwards DA. Testosterone, cortisol, and human competition. Horm Behav. 2016;82:21–37.

22. Björk J, Modig K, Kahn F, Ahlbom A. Revival of ecological studies during the COVID-19 pandemic. European Journal of Epidemiology. 2021;36(12):1225–9.

23. Pearce N. Epidemiology in a changing world: variation, causation and ubiquitous risk factors. Int J Epidemiol. 2011;40(2):503–12.

24. Nelson Mandela Foundation. Speech by Nelson Mandela at the Inaugural Laureus Lifetime Achievement Award, Monaco 2000: World Laureus Sports Awards Limited; 2000 [Available from: http://db.nelsonmandela.org/speeches/pub_view.asp?pg=item&ItemID=NMS1148.

